# Detection of new *Mycobacterium leprae* subtype in Bangladesh by genomic characterization to explore transmission patterns

**DOI:** 10.1101/2020.03.05.20031450

**Authors:** Maria Tió-Coma, Charlotte Avanzi, Els M. Verhard, Louise Pierneef, Anouk van Hooij, Andrej Benjak, Johan Chandra Roy, Marufa Khatun, Khorshed Alam, Paul Corstjens, Stewart T. Cole, Jan Hendrik Richardus, Annemieke Geluk

## Abstract

*Mycobacterium leprae*, the causative agent of leprosy, is an unculturable bacterium with a considerably reduced genome (3.27 Mb) compared to homologues mycobacteria from the same ancestry. *M. leprae* transmission is suggested to occur through aerosols but the exact mechanisms of infection remains unclear. In 2001, the genome of *M. leprae* was first described and subsequently four genotypes (1-4) and 16 subtypes (A-P) were identified providing means to study global transmission patterns for leprosy.

We investigated *M. leprae* carriage as well as infection in leprosy patients (n=60) and healthy household contacts (HHC; n=250) from Bangladesh using molecular detection of the bacterial element RLEP in nasal swabs (NS) and slit skin smears (SSS). In parallel, we explored bacterial strain diversity by whole-genome sequencing (WGS) and Sanger sequencing.

In the studied cohort in Bangladesh, *M. leprae* DNA was detected in 33.3% of NS and 22.2% of SSS of patients with bacillary index of 0 whilst in HHC 18.0% of NS and 12.3% of SSS were positive.

The majority of the *M. leprae* strains detected in this study belonged to genotype 1D (55%), followed by 1A (31%). Importantly, WGS allowed the identification of a new *M. leprae* genotype, designated 1B-Bangladesh (14%), which clustered separately between the 1A and 1B strains. Moreover, we established that the genotype previously designated 1C, is not an independent subtype but clusters within the 1D genotype.

Intraindividual differences were present between the *M. leprae* strains obtained including mutations in hypermutated genes, suggesting mixed colonization/infection or in-host evolution.

In summary, we observed that *M. leprae* is present in asymptomatic contacts of leprosy patients fueling the concept that these individuals contribute to the current intensity of transmission. Our data therefore emphasize the importance of sensitive and specific tools allowing post-exposure prophylaxis targeted at *M. leprae*-infected or -colonized individuals.

## Introduction

*Mycobacterium leprae* and the more recently discovered *Mycobacterium lepromatosis* (Han et al., 2008) are the causative agents of leprosy in humans as well as animals (Truman et al., 2011; Sharma et al., 2015; Avanzi et al., 2016; Honap et al., 2018; Schilling et al., 2019b; Tio-Coma et al., 2019). Leprosy is a complex infectious disease often resulting in severe, life-long disabilities and still poses a serious health threat in low-and middle income countries (WHO, 2019). Despite the very limited *M. leprae* genome variability (Singh and Cole, 2011), the disease presents with characteristically different clinico-pathological forms (Ridley and Jopling, 1966) due to genetically dependent differences in the immune response to the pathogen, resulting in the WHO classification from paucibacillary (PB) to multibacillary (MB) leprosy (Kumar et al., 2017). Notwithstanding the efficacy of multidrug therapy (MDT), approximately 210,000 new cases are still annually diagnosed and this incidence rate has been stable over the last decade (WHO, 2019). Aerosol transmission via respiratory routes is generally assumed to be the most probable way of bacterial dissemination (Bratschi et al., 2015; Araujo et al., 2016). Besides bacterial exposure other risk factors have been shown to be associated with development of leprosy such as genetic polymorphisms (Mira et al., 2004; Zhang et al., 2009; Wang et al., 2015; Sales-Marques et al., 2017), the clinical type of the leprosy index case within a household, immunosuppression (Moet et al., 2004), and nutritional factors (Dwivedi et al., 2019).

*M. leprae* is closely related to *Mycobacterium tuberculosis*, however, its genome has undergone a reductive evolution resulting in a genome of only 3.27 Mb compared to the 4.41 Mb of *M. tuberculosis*’ (Cole et al., 2001). Part of the genes lost in *M. leprae* included vital metabolic activity, causing it to be an obligate intracellular pathogen which cannot be cultured in axenic media that requires support of a host to survive. This poses major limitations to obtain sufficient bacterial DNA for research purposes including whole genome sequencing (WGS). Nevertheless, in 2001 the genome of *M. leprae* was first published (Cole et al., 2001) leading to the classification of *M. leprae* into four main genotypes (1-4) (Monot et al., 2005) and subsequently further allocation into 16 subtypes (A-P) (Monot et al., 2009; Truman et al., 2011). The genome of *M. leprae* contains several repetitive elements such as RLEP which present 37 copies and has been widely applied in molecular diagnostics to specifically detect the presence of this mycobacterium (Donoghue et al., 2001; Truman et al., 2008; Martinez et al., 2011; Braet et al., 2018).

Single-nucleotide polymorphisms (SNP) genotyping and WGS are powerful approaches to investigate pathogen transmission as well as bacterial dissemination and evolution through genome characterization (Monot et al., 2005; Monot et al., 2009; Han and Silva, 2014). The limited variation observed in the *M. leprae* genome permits the reconstruction of historic human migration patterns and the origin of *M. leprae* (Donoghue, 2019). Over the years, several studies have contributed to the detection and characterization of *M. leprae* genomes originating from patients all around the world (Monot et al., 2005; Monot et al., 2009; Benjak et al., 2018) as well as from ancient skeletons (Suzuki et al., 2010; Schuenemann et al., 2013; Mendum et al., 2014; Krause-Kyora et al., 2018; Schuenemann et al., 2018), red squirrels (Avanzi et al., 2016; Schilling et al., 2019a; Tio-Coma et al., 2019), armadillos (Truman et al., 2011; Sharma et al., 2015), non-human primates (Honap et al., 2018) and soil (Lavania et al., 2006; Lavania et al., 2008; Turankar et al., 2012; Turankar et al., 2014; Turankar et al., 2016; Tió-Coma et al., 2019; Turankar et al., 2019). Moreover, skeleton remains have been successfully applied to retrospectively assess whether individuals who contributed to the care of leprosy patients such as the priest Petrus Donders, had developed leprosy (Van Dissel et al., 2019). In the last few years, new tools were developed allowing direct sequencing of *M. leprae* from various types of clinical isolates (Avanzi et al., 2016; Benjak et al., 2018; Schuenemann et al., 2018). However, these methods were never applied on challenging samples such as slit skin smears (SSS) and nasal swabs (NS) containing a low amount of bacterial DNA compared to skin lesions of patients.

Household contacts of leprosy patients are a high risk group for developing the disease (Richardus et al., 2019), and might serve as asymptomatic carriers contributing to bacterial dissemination. PCR and quantitative PCR (qPCR) are reliable techniques to detect *M. leprae* DNA and have been proposed as tools for early diagnosis of leprosy, particularly among household contacts of newly diagnosed patients (Gama et al., 2018; Gama et al., 2019). In Brazil, *M. leprae* DNA has been detected in 15.9% of healthy household contacts (HHC) in SSS, 9.7% in blood (Gama et al., 2018) and 8.9 to 49.0% in NS (Brito e Cabral et al., 2013; Araujo et al., 2016; Carvalho et al., 2018). Other studies from India, Indonesia and Colombia reported 21% of *M. leprae* positivity in SSS of HHC (Turankar et al., 2014), 7.8% (van Beers et al., 1994) and 16.0% in NS (Romero-Montoya et al., 2017).

Detection of host markers, such as serum IgM levels of anti-*M. leprae* phenolic glycolipid I (PGL-I), represents an alternative approach to diagnose infected individuals (Penna et al., 2016; van Hooij et al., 2017; Barbieri et al., 2019). However, although detection of *M. leprae* DNA as well as antibodies against PGL-I indicate infection with *M. leprae*, this does not necessarily result in disease. Thus, these tests alone are not sufficient to identify the complete leprosy spectrum (Spencer and Brennan, 2011; van Hooij et al., 2019).

Bangladesh is a leprosy endemic country reporting up to 3,729 new leprosy cases in 2018 (WHO, 2019). However, *M. leprae* whole genomes (n=4) from Bangladesh, have only been described in one study (Monot et al., 2009) in which genotypes 1A, 1C and 1D were identified. To gain more insight into *M. leprae* genome variation and transmission routes in endemic areas in Bangladesh as well as the potential role of asymptomatic carriers, we further explored the diversity and transmission of *M. leprae* in four districts of the northwest of Bangladesh. We collected SSS and NS of 31 leprosy patients with a high bacterial load as well as 279 of their household contacts and characterized *M. leprae* DNA by WGS or Sanger sequencing. The resulting genotypes were correlated to the subjects’ GIS location. Additionally, this is the first study to examine *M. leprae* DNA detection in comparison to anti-PGL-I IgM levels in plasma measured by up-converting phosphor lateral flow assays (UCP-LFAs).

## Materials and methods

### Study design and sample collection

Newly diagnosed leprosy patients (index case, n=31) with bacteriological index (BI) ≥ 2 and 3-15 household contacts of each index case (n=279) were recruited between July 2017 and May 2018 (Table S1, Supplementary Data 1) in four districts of Bangladesh (Nilphamari, Rangpur, Panchagar and Thakurgaon). Patients with five or fewer skin lesions and BI 0 were grouped as PB leprosy. Patients with more than five skin lesions were grouped as MB leprosy and BI was determined. The prevalence in the districts where this study was performed was 0.9 per 10,000 and the new case detection rate 1.18 per 10,000 (Rural health program, the leprosy mission Bangladesh, yearly district activity report 2018). For *M. leprae* detection and characterization, SSS from 2-3 sites of the earlobe and NS (tip wrapped with traditional fiber, CLASSIQSwabs, Copan, Brescia, Italy) were collected and stored in 1 ml 70% ethanol at −20 °C until further use. For immunological analysis, plasma was collected (van Hooij et al., 2017; van Hooij et al., 2018; van Hooij et al., 2019).

Subjects included in the study were followed up for surveillance of new case occurrence for ≥ 24 months after sample collection.

### Ethics Statement

Subjects were recruited following the Helskinki Declaration (2008 revision). The National Research Ethics Committee approved the study (BMRC/NREC/2016-2019/214) and participants were informed about the study objectives, the samples and their right to refuse to take part or withdraw without consequences for their treatment. All subjects gave informed consent before enrollment and treatment was provided according to national guidelines.

### DNA isolation from slit skin smears and nasal swabs

DNA was isolated using DNeasy Blood & Tissue Kit (Qiagen, Valencia, CA) as per manufacturer’s instructions with minor modifications. Briefly, tubes containing 1 ml 70% ethanol and SSS were vortexed for 15 seconds. SSS were removed and tubes were centrifuged for 15 minutes at 14000 rpm. Supernatants were removed and buffer ATL (200 μl) and proteinase K (20 μl) added. NS were transferred to new microtubes and the microtubes containing the remaining ethanol were centrifuged at 14000 rpm for 15 minutes. Supernatants were removed and NS were inserted again in the tubes, prior addition of ATL buffer (400 μl) and proteinase K (20 μl). SSS and NS samples were incubated at 56 °C for 1 h at 1100 rpm. Next, AL buffer (200 μl) was added and incubated at 70 °C for 10 min at 1400 rpm. Column extraction was performed after absolute ethanol precipitation (200 μl) as per manufacturer’s instructions. To avoid cross contamination tweezers were cleaned first with hydrogen peroxide and then with ethanol between samples.

### RLEP PCR and qPCR

RLEP PCR (Donoghue et al., 2001) was performed as previously described (Tió-Coma et al., 2019). Briefly, the 129 bp RLEP sequence was amplified in 50 µl by addition of 10 µl 5x Gotaq® Flexi buffer (Promega, Madison, WI), 5 µl MgCl_2_ (25 mM), 2 µl dNTP mix (5 mM), 0.25 µl Gotaq® G2 Flexi DNA Polymerase (5 u/µl), 5 µl (2 µM) forward and reverse primers (Table S2) and 5 µl template DNA, water (negative control) or *M. leprae* DNA (Br4923 or Thai-53 DNA, BEI Resources, Manassas, VA) as positive control. PCR mixes were subjected to 2 min at 95 °C followed by 40 cycles of 30 s at 95 °C, 30 s at 65°C and 30 s at 72 °C and a final extension of 10 min at 72 °C. PCR products (15µl) were used for electrophoresis in a 3.5% agarose gel at 130V. Amplified DNA was visualized by Midori Green Advance staining (Nippon Genetics Europe, Dueren, Germany) using iBright^™^ FL1000 Imaging System (Invitrogen, Carlsbad, CA).

Samples from index cases and a selectin of contacts for sequencing were also evaluated by qPCR (Martinez et al., 2009). The mix included 12.5 µl TaqMan Universal Master Mix II (Applied Biosystems, Foster City, CA), 0.5 µl (25 µM) forward and reverse primers (Table S2), 0.5 µl (10 µM) TaqMan probe (Table S2) and 5 µl template DNA were mixed in a final volume of 25 µl. DNA was amplified using the following profile: 2 min at 50°C and 10 min at 95°C followed by 40 cycles of 15 s at 95°C and 1 min at 60°C with a QuantStudio 6 Flex Real-Time PCR System (Applied Biosystems). Presence of *M. leprae* DNA was considered if a sample was positive for RLEP qPCR with a cycle threshold (Ct) lower than 37.5 or was positive for RLEP PCR at least in two out of three indecently performed PCRs to avoid false positives.

### Library preparation and enrichment

A total of 60 DNA extracts were selected for sequencing, including 30 from SSS and 30 from NS (Figure S1, Supplementary Data 1). At least one sample from each index leprosy patient was selected as well as RLEP positive samples of HHC or patients who were household contacts of the index case (selection based on Ct value and household overlap). A maximum of 1µg of DNA in a final volume of 50µL was mechanically fragmented to 300 bp using the S220 Focused-ultrasonicator (Covaris) following the manufacturer’s recommendations and cleaned-up using a 1.8x ratio of AMPure beads. Up to 1µg of fragmented DNA was used to prepare indexed libraries using the Kapa Hyperprep kit (Roche) and the Kapa dual-indexed adapter kit as previously described (Benjak et al., 2018) followed by two rounds of amplification. All libraries were quantified using the Qubit fluorimeter (Thermo Fisher Scientific, Waltham, MA), and the fragment size distribution was assessed using a fragment analyzer.

Libraries were target enriched for the *M. leprae* genome using a custom MYbaits Whole Genome Enrichment kit (ArborBioscence) as previously described (Honap et al., 2018). Briefly, biotinylated RNA baits were prepared using DNA from *M. leprae* Br4923. A total of 1500 ng of each amplified libraries was used for enrichment. Each library was pooled prior to enrichment with another library with similar qPCR Ct value. Enrichment was conducted according to the MYbaits protocol with the hybridization being carried out at 65 °C for 24 hours. After elution, all pools were amplified using the Kapa amplification kit with universal P5 and P7 primers (Roche). All amplification reactions were cleaned up using the AMPure beads (1X ratio).

### Illumina sequencing

Pools were multiplexed on one lane of a NextSeq instrument with a total amount of 20-30 million reads per pools. Some libraries were deep sequenced based on the mapping statistics obtained in the first run. Raw reads were processed and aligned to *M. leprae* TN reference genome (GenBank accession number AL450380.1) as previously described using an in-house pipeline (Benjak et al., 2018). A minimum depth coverage of 5 was considered for further phylogenetic analysis.

### Sequencing analysis

Genome comparison was based on analysis of SNPs (analyzed with VarScan v2.3.9(Koboldt et al., 2012)) and Indels (analyzed with Platypus v0.8.171(Rimmer et al., 2014)) as formerly reported (Benjak et al., 2018). The newly sequenced *M. leprae* genomes were aligned with 232 genomes available in public databases (Schuenemann et al., 2018; Avanzi et al., In press). Sites below 90 and above 10% alignment difference were also reported. A comparison to 259 *M. leprae* genomes (including 27 new genomes) allowed the identification of unique SNPs per index case. Each candidate SNP or Indel was checked manually on Integrative Genomics Viewer (Robinson et al., 2011).

### Genotyping and antimicrobial resistance by Sanger sequencing

To further characterize the *M. leprae* strains for which the whole genome sequence was not obtained, specific primers were designed to perform Sanger sequencing based on unique SNPs (Table S3 and S4) of each index case strain. Additionally, Sanger sequencing was performed after amplifying several loci (Table S2) to subtype the genomes based on standard the *M. leprae* classification (Monot et al., 2009; Truman et al., 2011) and to determine antimicrobial resistance to rifampicin (*rpoB*), dapsone (*folP1*) or ofloxacin (*gyrA*). PCRs were performed with 5 µl of template DNA using the aforementioned PCR mixes. DNA was denatured for 2 minutes at 95°C, followed by 45 cycles of 30 s at 95°C, 30 s at 50-58 °C and 30 s at 72 °C and a final extension cycle of 10 min at 72°C. PCR products were resolved by agarose gel electrophoresis as explained above. PCR products showing a band were purified prior to sequencing using the Wizard SV Gel and PCR Clean-Up System (Promega). Sequencing was performed on the ABI3730xl system (Applied Biosystems) using the BigDye Terminator Cycle Sequencing Kit (Thermo Fisher Scientific). Sequences were analyzed using Bioedit v7.0.5.3.

### Anti-PGL-I UCP-LFA

UCP-LFAs were performed using the LUMC developed LFA based on luminescent up-converting reporter particles for quantitative detection of anti-*M. leprae* PGL-I IgM as previously described (van Hooij et al., 2017; van Hooij et al., 2018; van Hooij et al., 2019). Plasma samples (n=308, 2 samples excluded due to labeling mistake) were thawed and diluted (1:50) in assay buffer. Strips were placed in microtiter plate wells containing 50 µl diluted samples and target specific UCP conjugate (PGL-I, 400 ng). Immunochromatography continued for at least 30 min until dry. Scanning of the LFA strips was performed by LFA strip readers adapted for measurement of the UCP label (UPCON; Labrox, Finland). Results are displayed as the Ratio (R) value between Test and Flow-Control signal based on relative fluorescence units (RFUs) measured at the respective lines. The threshold for positivity for the αPGL-I UCP-LFA was 0.10.

## Results

### *M. leprae* detection in patients and healthy household contacts

At diagnosis of the index cases and recruitment of contacts into this study 250 household contacts had no signs or symptoms of leprosy or other diseases (HHC), whereas 22 household contacts were diagnosed as PB and seven as MB patients (Table S1, Supplementary Data 1).

Presence of *M. leprae* DNA was determined by RLEP PCR or qPCR in SSS and NS of leprosy patients and HHC (Figure 1, Supplementary Data 1): as expected in MB patients with BI 2-6 *M. leprae* DNA was almost always detectable in both SSS (96.8%) and NS (90.9%). This was much lower in PB and MB patients with BI 0 ranging from 22.2% in SSS to 33.3% in NS. Positivity rates in HHC were not very different from those observed for PB and MB patients with BI 0, with 12.3% positive samples in SSS and 18.0% in NS. Moreover, the overall Ct range was lower for SSS [16.3-37.1] compared to NS [20.1-39.4] showing that SSS contained more *M. leprae* DNA and is a preferred sample for its detection (Supplementary Data 1).

**Figure 1.**
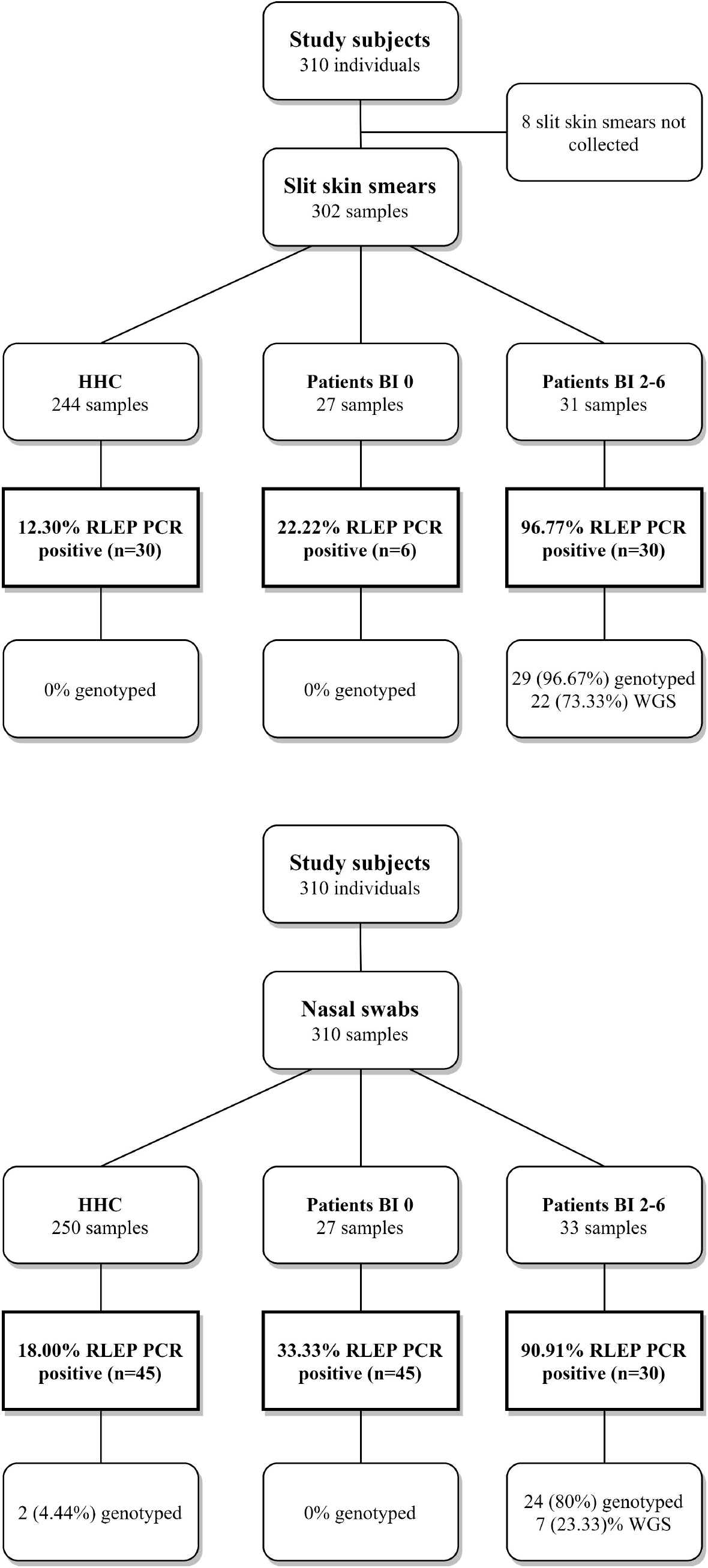
Study design, RLEP positivity and genotyped samples. Flow diagram providing an overview of the subjects recruited for this study. Slit skin smears (SSS) and nasal swabs (NS) collected per group; healthy household contacts (HHC), paucibacillary (PB) or multibacillary (MB) patients with BI 0, and MB patients with a bacteriological index (BI) 2-6. MB patients with BI 1 were not diagnosed within the course of this study. DNA was isolated from SSS and NS and screened for *M. leprae* DNA by RLEP PCR. Samples were genotyped by Sanger sequencing (Monot et al., 2009; Truman et al., 2011) or Whole Genome Sequencing (Benjak et al., 2018). Percentages of the samples positive for RLEP PCR and genotyped are shown.

HHC (n=250) were followed up clinically for ≥ 24 months after sample collection and four of them developed leprosy within the first year. RLEP PCR performed on DNA isolated before disease occurrence showed a positive result from SSS in one patient (5 months before diagnosis) and a positive result from NS in another (8 months before diagnosis). All of the new cases developed PB leprosy with BI of 0 and three were genetically related to the index case (parent and child of index case H03 and second degree relative of index case H30) and one was the spouse (index case H10).

### Genome typing and antimicrobial resistance

*M. leprae* genomes of SSS and NS were genotyped by WGS or Sanger sequencing. A total of 60 samples (30 SSS and 30 NS) were selected for WGS with an RLEP qPCR Ct ranging from 16.2 to 37.2 (Figure S1, Supplementary Data 1). A total of 27 samples from 21 subjects (21 SSS and 6 NS) were successfully sequenced with a coverage ≥ 5 (Table S5). The limiting Ct value was 26.2 for SSS and 24.2 for NS.

On applying the genotyping system described by (Monot et al., 2009; Truman et al., 2011), the following genotypes were found for these 21 subjects: 1A (n=5), 1B (n=4), 1C (n=3) and 1D (n=9). Interestingly, the four newly sequenced 1B genotype strains do not cluster with the two previously described 1B strains from Yemen and Martinique (Figure 2). Instead, they form a new cluster in the phylogenetic tree located between genotypes the 1A and 1B, which we refer to as 1B-Bangladesh (Figure 2, blue, Supplementary Data 1). Using Sanger sequencing, the *M. leprae* strain for eight additional individuals were determined as 1A (n=4) or 1D (n=4). Three subjects carried genotype 1 but subtype could not be established (Supplementary Data 1).

**Figure 2.**
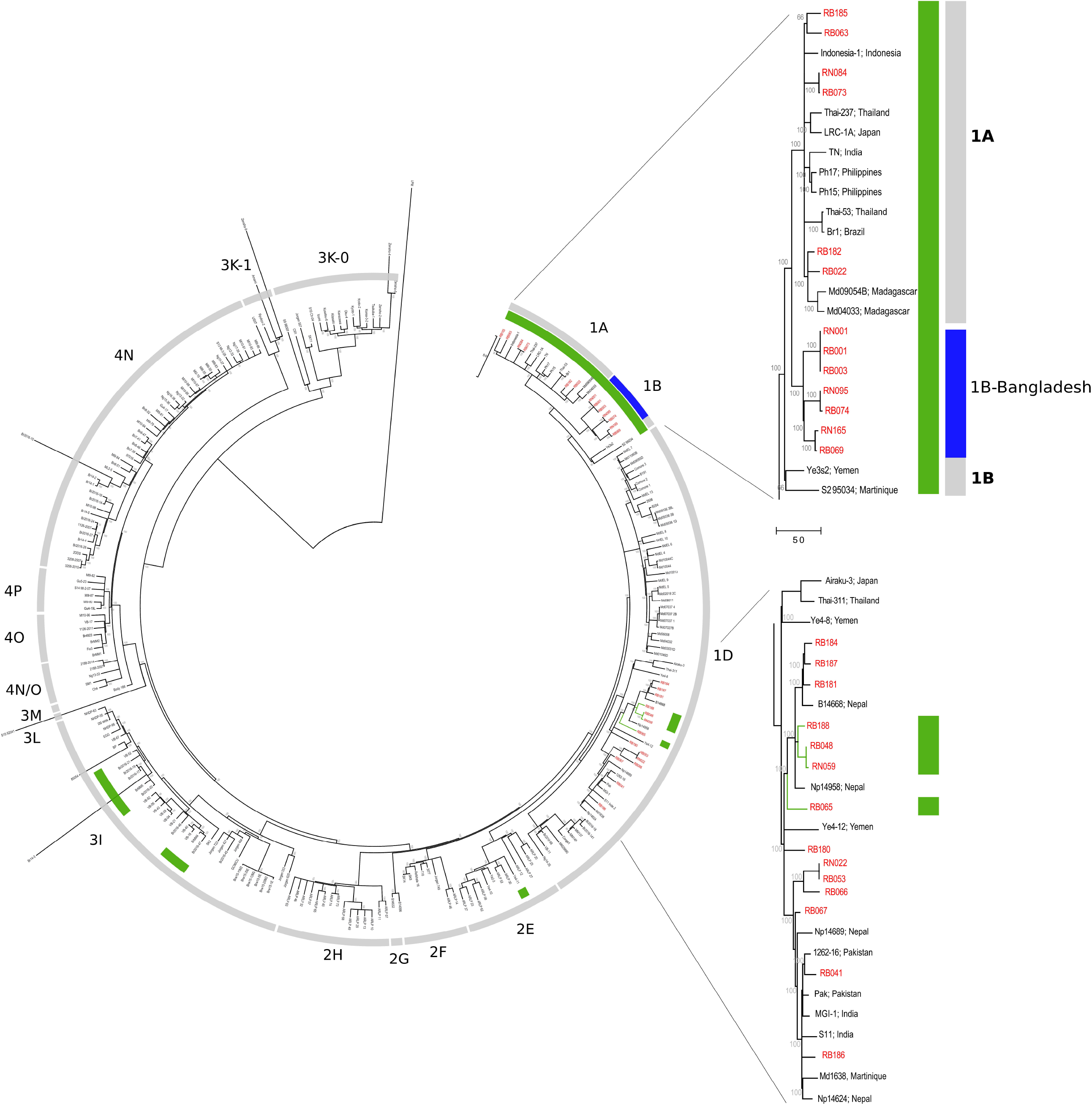
Phylogeography of *M. leprae* strains. Maximum parsimony tree of 259 genomes of *M. leprae* built in MEGA 7. Support values were obtained by bootstrapping 500 replicates. Branch lengths are proportional to nucleotide substitutions. The tree is rooted using *M. lepromatosis*. The strains from Bangladesh are shown in red and their exact organization in the tree is shown in the two zoomed sections of the genotypes 1A-B and 1D. Strains with an A at SNP61425 in the *esxA* gene are shown in green. The specific 1B-Bangladesh genotype/cluster of Bangladesh strains is shown in blue.

The SNP used to differentiate genotype 1C (A61425G; Met90Thr, mutated in genotypes 1D and 2-4) is located at *esxA*. In contrast to previous observations (Monot et al., 2009; Truman et al., 2011), we found that this position is not phylogenetically informative as it is also found unmutated (A; Met) in strains from the genotype 3I and 2E (Figure 2, green, Supplementary Data 2). Moreover, the 1C strains clustered in the middle of the 1D group suggesting that the previously described genotype 1C is part of the 1D genotype.

Finally, antimicrobial resistance was assessed in all genotyped strains either by WGS or Sanger sequencing. The latter was successful on 18 samples for *rpoB*, five sample for *folP1* and 15 samples for *gyrA* (Supplementary Data 1). None of the strains with a complete genome harbored drug-resistance mutations. One NS sample containing a missense mutation in the *rpoB* gene (Ser456Thr) in 50% of the sequences potentially leading to antimicrobial resistance (2017) was identified by Sanger sequencing. Moreover, although not causing resistance, up to two silent mutations in three different positions of the *rpoB* gene relevant for antimicrobial resistance (432, 441 and 456) were also observed in several subjects.

### Distribution and possible transmission of *M. leprae* genotypes

The most prevalent *M. leprae* genotype in the studied area of Bangladesh is 1D, found in 55% of the individuals (n=16, Table 1, Supplementary Data 1), followed by 1A in 31% (n=9), and 1B-Bangladesh in 14% (n=4). Genotype 1D is the most widely distributed throughout the whole area studied (Figure 3, blue and purple), whilst genotypes 1A and the here identified genotype 1B-Bangladesh are only observed in the eastern area (green and orange respectively). The latter genotype was found in 4 individuals: two from the same household and two unrelated subjects residing 56, 51 and 11 km from each other. However, due to privacy regulations on patient information to third parties it could not be established whether subjects in different households had had contact with any of the others.

**Table 1.** *M. leprae* genotypes identified in Bangladesh.

| Genotype | Number of individual | % |
| --- | --- | --- |
| 1A | 9 | 31.0 |
| 1B-Bangladesh | 4 | 13.8 |
| 1D | 13 | 44.8 |
| 1D-*esxA* | 3 | 10.4 |
| 1* | 3 |  |

**Figure 3.**
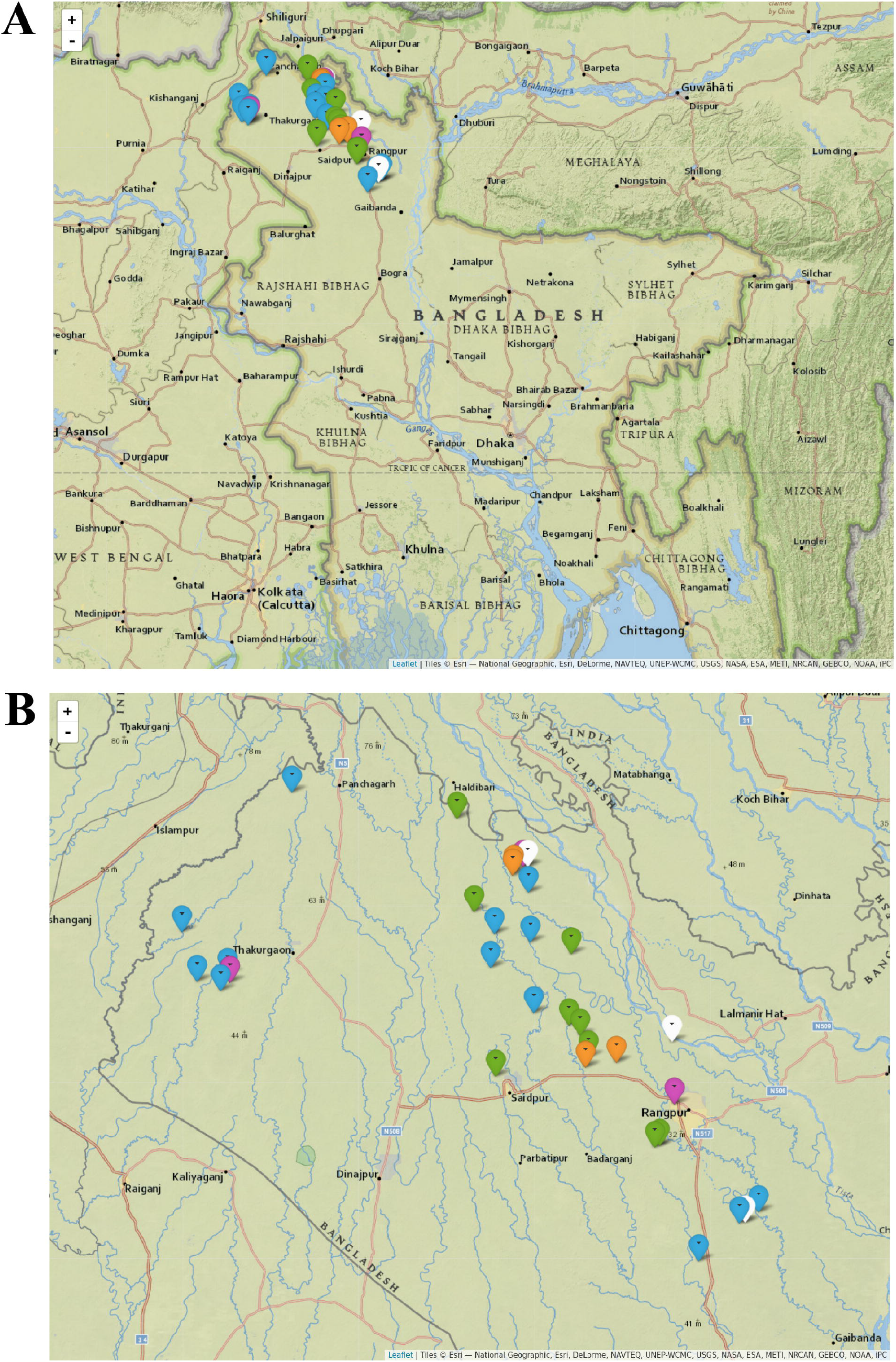
Distribution of *M. leprae* genotypes in Bangladesh. Map of Bangladesh including markers indicating the residence of every subject with at least one sample genotyped for *M. leprae* (A), and zoomed into the area of interest (B). Each marker indicates an individual for whom *M. leprae* genotype was determined, either from slit skin smear, nasal swab or both samples. Genotype 1A is shown in green, 1B-Bangladesh in orange, 1D in blue, 1D-*esxA* in purple and 1* in white. 1D-*esxA* is 1D subtype containing an A at SNP61425 in the *esxA* gene, formerly grouped as 1C (Monot et al., 2009; Truman et al., 2011). 1* are samples with genotype 1 for which the subtype could not be determined. The figure was drawn in R (v3.4.3) with the package *leaflet* (v2.0.2) using maps from Esri – National Geographic with permission. Scale Not Given. “National Geographic World Map”. December 13, 2011. http://www.arcgis.com/home/item.html?id=b9b1b422198944fbbd5250b3241691b6 (September 2, 2019).

In a total of four households the same *M. leprae* genotype was detected in two individuals (Supplementary Data 1). In the first household, both subjects were MB patients and WGS showed no genetic variation at all between both patients’ genomes (RB001 and RB003, 1B-Bangladesh genotype, Supplementary Data 2). In the second household with two MB patients, the *M. leprae* whole genome was only obtained from the index case but the same genotype, 1A, and a strain-specific SNP of the index case (Table S3 and S4) was also identified by Sanger sequencing in the other patient (RB182 and RB266). In the last two households, the genotype of strains from both MB index cases’ were determined by WGS (RB030, genotype 1D) and, by Sanger sequencing (RB065, genotype 1D-*esxA*), while the *M. leprae* genotype 1 was located in the NS of both HHC but no further subtyping was possible.

### Comparison of *M. leprae* genomes from SSS and NS

*M. leprae* whole genomes of six patients were successfully recovered from both SSS and NS. Genomic comparison showed no differences between DNA from SSS and NS for two patients: RB001-RN001 (genotype 1B-Bangladesh) and RB048-RN059 (genotype 1D-*esxA*, Supplementary Data 2, Figure 2). In a third patient (RB073-RN084, genotype 1A), both strains were identical except that in the NS strain 13% of 32 reads in *ml1512* harbored a T1824441C (Gly56Asp) (Table 2). Interestingly, *ml1512* which encodes a ribonuclease J is one of the most mutated genes among all *M. leprae* strains (Benjak et al., 2018) and mutations at this gene were also observed in two different patients: in the NS of RN022-RB053 (genotype 1D) 28% of 115 reads had a mutated allele (G1823127A; Ser494Leu) and 8.5% of 59 reads had an insertion of a C at position 1823614 probably leading to a deleterious frameshift; in the SSS of RB074-RN095 (genotype 1B-Bangladesh) 91% of 158 reads presented a missense mutation (G1823098A; Leu504Phe). Interestingly, RB074 harbored a G660474C mutation in *metK*, a probable methionine adenosyl-transferase, which was also found in 76% of 16 reads of the NS and is uniquely found in this subject’s *M. leprae* genomes. Additionally, RN095 also displayed mutations at several positions in *ml1750* (a putative nucleotide cyclase): 60% of 40 reads had C2116695A (Pro100Thr), 23% of 40 reads had A2116670G (Gln108Arg) and 26% of 27 reads had G2116670A mutation (Arg168His). These positions were partially or totally mutated in other strains from different genotypes: SM1 (100% Pro100Ser; genotype 4), Ml9-81 (Mali, 30% Arg168His; genotype 4N) and Md05036 (Madagascar, 90% Gln108Arg, genotype 1D-Malagasy) (Benjak et al., 2018; Avanzi et al., In press).

**Table 2.** Intraindividual *M. leprae* genomic differences.

| Samples | Mutation | Gene | % reads | Aligned | % reads | Aligned |
| --- | --- | --- | --- | --- | --- | --- |
|  |  |  | SSS | reads SSS | NS | reads NS |
| **RB073-RN084** | T1824441C; Gly56Asp | *ml1512* |  |  | 13% | 32 |
| **RB053-RN022** | G1823127A; Ser494Leu | *ml1512* |  |  | 28% | 115 |
|  | 1823614_1823615insC | *ml1512* |  |  | 8.5% | 59 |
| **RB074-RN095** | G1823098A; Leu504Phe | *ml1512* | 91% | 158 |  |  |
|  | G660474C; Val252Leu | *metK* | 100% | 196 | 76% | 16 |
|  | C2116695A; Pro100Thr | *ml1750* |  |  | 60% | 40 |
|  | A2116670G; Gln108Arg | *ml1750* |  |  | 23% | 40 |
|  | G2116695A; Arg168His | *ml1750* |  |  | 26% | 27 |
| **RB069-RN165** | C9231T; Leu34Phe | *glpQ* |  |  | 25% | 257 |
|  | C2121552T; Val226Ile | *ml1752* |  |  | 16% | 307 |

2 Genomic differences between *M. leprae* genomes obtained from slit skin smears (SSS) and nasal swabs 3 (NS) of the same MB patient. Percentage of mutated reads and total number of reads aligned at the 4 position of the mutation. No differences were found between the SSS and NS genomes of two patients: 5 RB001-RN001 and RB048-RN059.

The patient with the *M. leprae* strains that were the most genetically different between the NS and SSS carried the genotype 1B-Bangladesh (RB069 and RN165). The NS strain had a mixed population in *glpQ* (25% of 257 reads C9231T, Leu34Phe) and *ml1752* (16% of 307 reads C2121552T, Val226Ile). These genes encode a glycerophosphoryl diester phosphodiesterase and a conserved hypothetical protein. Notably, *ml1752* is also one of the most hypermutated genes in *M. leprae* (Benjak et al., 2018). For 11 patients a whole genome sequence was recovered only from SSS but Sanger sequencing was successfully performed to identify the subtype in NS. The same subtype observed in SSS was also found in the NS of these 11 patients. Moreover, unique *M. leprae* SNPs identified in the genomes of the SSS (Table S3 and S4) were also detected in seven of the genomes of the NS of these patients (Supplementary Data 1).

### Combining host and pathogen detection

Anti-PGL-I IgM levels were determined in plasma of 308 subjects. All MB patients with BI 2-6 (n=33) showed high levels for anti-PGL-I IgM (Table 3) in line with the general consensus (Geluk et al., 2011; van Hooij et al., 2017). Out of the patients (both MB and PB) with BI 0 (n=27), nine (33.3%) were positive for anti-PGL-I IgM. Similarly, 36.8% of HHC showed positivity (n=92). From these 92 positive individuals, 70 were neither positive for SSS nor NS RLEP PCR (Supplementary Data 1). Of the four contacts who developed leprosy within the first year after sample collection, two were positive for anti-PGL-I IgM whilst negative for RLEP PCRs 10 and 12 months before diagnosis. Since the two other subjects had a positive RLEP PCR in SSS or NS 5 or 8 months before diagnosis, it can be concluded that all of the new cases showed positivity either for host-or pathogen-associated diagnostics 5-12 months before developing disease.

**Table 3.**
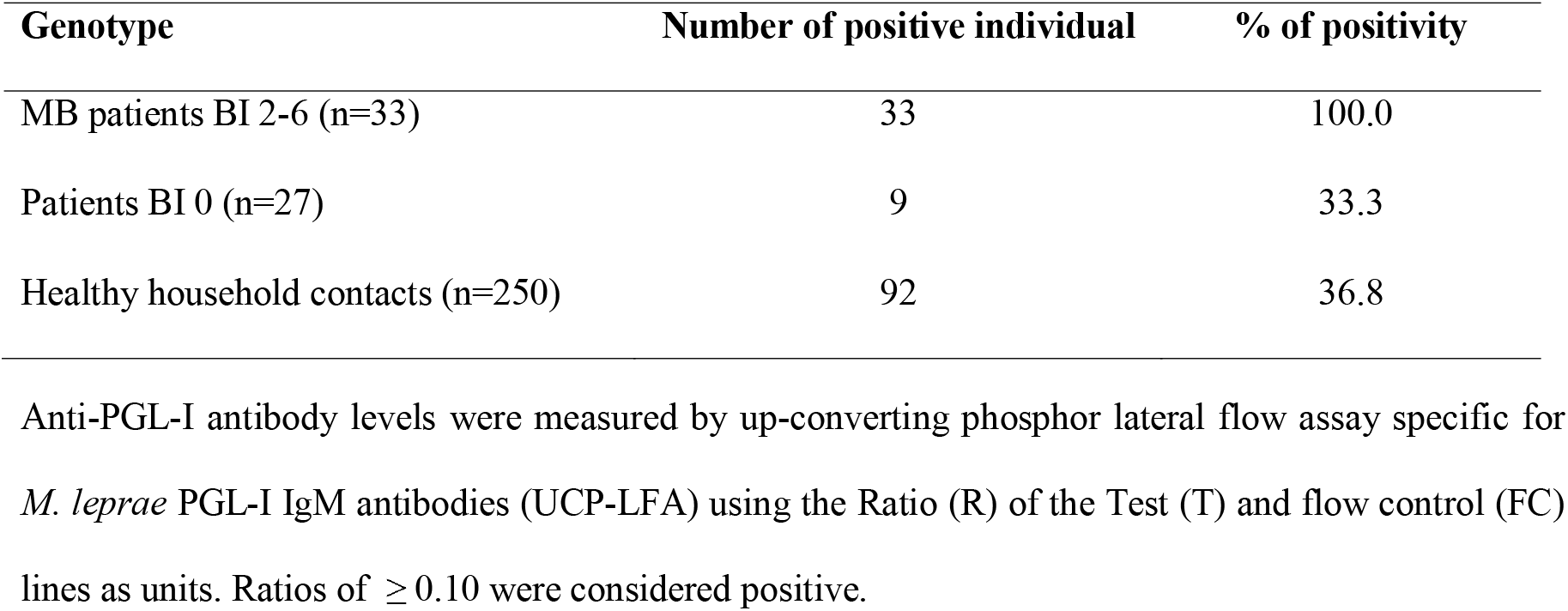
Anti-PGL-I IgM positivity.

| Genotype | Number of positive individual | % of positivity |
| --- | --- | --- |
| MB patients BI 2-6 (n=33) | 33 | 100.0 |
| Patients BI 0 (n=27) | 9 | 33.3 |
| Healthy household contacts (n=250) | 92 | 36.8 |

2 Anti-PGL-I antibody levels were measured by up-converting phosphor lateral flow assay specific for 3 *M. leprae* PGL-I IgM antibodies (UCP-LFA) using the Ratio (R) of the Test (T) and flow control (FC) 4 lines as units. Ratios of $\geq 0.10$ were considered positive.

Individual anti-PGL-I levels were compared to RLEP Ct values in SSS and NS samples (Figure 4), showing an expected negative correlation between anti-PGL-I ratio and Ct value since both values are associated with BI. A subtle difference can be observed in the correlation between anti-PGL-I IgM levels and RLEP Ct if the qPCR was performed on either SSS or NS DNA, with a coefficient of determination (R^2^) 0.73 and 0.69 respectively.

**Figure 4.**
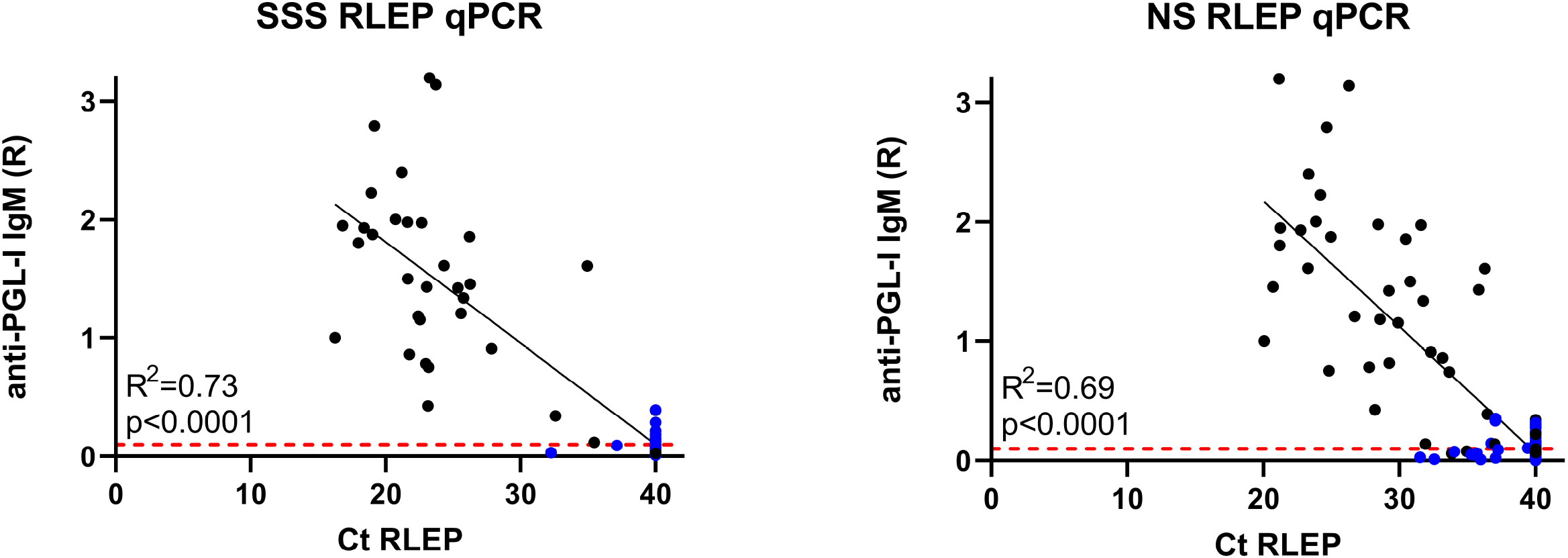
Correlation of IgM antibodies against PGL-I to Ct of RLEP qPCR. Quantified levels of pathogen DNA (qPCR) and host immunity were correlated for samples selected for qPCR analysis based on RLEP positivity in multiple individuals in one household. Each dot represents a sample from one individual; leprosy patients are indicated in black, and healthy household contact in blue. Anti-PGL-I antibody levels were measured by up-converting reporter particles lateral flow assay specific for *M. leprae* PGL-I IgM antibodies (αPGL-I UCP-LFA) using the Ratio (R) of the Test (T) and flow control (FC) lines as units. Ratios of ≥ 0.10 were considered positive as indicated by the red dashed line. RLEP cycle threshold (Ct) values are indicated on the x-axis and were measured by qPCR to detect *M. leprae* DNA in slit skin smears (SSS, left) and nasal swabs (NS, left). Undetermined Cts are depicted as Ct 40.

## Discussion

In this study we investigated *M. leprae* transmission patterns in Bangladesh by detecting and sequencing *M. leprae* DNA derived from SSS and NS of patients and their household members. Our data represents the first report of *M. leprae* DNA detection in HHC from Bangladesh. We observed moderate positivity in HHC which was similar to positivity of leprosy patients with BI 0. A new genotype, 1B-Bangladesh, was sequenced and we showed that the previously described 1C genotype is part of the 1D group. Additionally, a negative correlation between RLEP Ct values indicating the amount of *M. leprae* DNA and anti-PGL-I IgM levels was observed.

*M. leprae* DNA detection frequency in HHC from Bangladesh (12.3% in SSS and 18.0% in NS) was in line with previous studies conducted in several hyperendemic areas of Brazil, Colombia and Indonesia (van Beers et al., 1994; Brito e Cabral et al., 2013; Romero-Montoya et al., 2017; Carvalho et al., 2018; Gama et al., 2018). In India higher positivity (21%) in SSS of HHC was reported (Turankar et al., 2014) whereas in a Brazilian study from Uberlandia, up to 49% of positivity in NS was observed (Araujo et al., 2016). Three factors may limit the translation of these high positive results from India and Brazil to our study: i) the sample sizes of the Indian and Brazilian studies were smaller (n=28 and n=104, respectively versus n=250 HHC in this study); ii) we conducted a more stringent approach by testing the samples in three independent PCRs; and iii) the epidemiology and incidence of MB cases in India and Brazil differ from the studied area in Bangladesh where MB leprosy cases occur less frequently than PB and also usually display a low BI (Richardus et al., 2017).

*M. leprae* DNA in the nose does not indicate disease but (transient) colonization whilst presence of *M. leprae* in SSS indicates infection. Thus, the higher RLEP PCR positivity in NS compared to SSS in patients with BI 0 and HHC likely represents the (virtual) absence of bacteria causing infection in these individuals despite colonization.

A longitudinal study conducted in Brazil (Manta et al., 2019), investigated SSS from 995 HHC by qPCR including follow-up for at least 3 years with occurrence of five new cases. The authors reported 20% qPCR positivity in HHC representing future new cases compared to 9% in HHC without disease. However, this difference was not significant. In line with that study, we found that *M. leprae* DNA detection was slightly higher (25% vs 18% in NS and 25% vs 12% in SSS) in contacts who developed disease compared to those who did not. Additionally, we determined anti-PGL-I IgM levels, which correlated well with Ct qPCR values. Notwithstanding this correlation, serology provided added value: when positivity in any of the three techniques was considered (NS PCR, SSS PCR or anti PGL-I), all of the contacts (n=4) who developed leprosy within the first year after sample collection, were identified. In agreement with this, a combination of host and pathogen markers was previously integrated in a machine learning model using qPCR and serological data (antibodies against LID-1 or ND-O-LID) (Gama et al., 2019) to identify prospective leprosy patients among contacts leading to an increased sensitivity in diagnosis, particularly in PB leprosy. It is of note that in our study, three of the four contacts who developed leprosy were genetically related to the index cases in their households, stressing the previously described role of genetic inheritance in the development of leprosy (Mira et al., 2004; Zhang et al., 2009; Wang et al., 2015; Sales-Marques et al., 2017; Uaska Sartori et al., 2020). For this reason, the association between leprosy and the genetics of this Bangladeshi population is currently being studied.

Genotype 1 was identified in all the *M. leprae* genomes retrieved from Bangladesh, consistent with previous data from (Monot et al., 2009). In Bangladesh, leprosy was likely introduced through the southern Asian route (genotype 1) leading to the spread of *M. leprae* into the Indian subcontinent, Indonesia and the Philippines (Monot et al., 2009; Benjak et al., 2018). Subtype 1D was predominantly present in Bangladesh but in addition we detected 1A and identified a new 1B-Bangladesh genotype. This new genotype is thus far restricted to Bangladesh and two of the four individuals carrying this strain were part of the same household whilst the other two did not have any relationship with each other and were located in different areas with a distance of up to 56 km between them. This suggests that this new genotype could be a common subtype in Bangladesh although additional studies are required to confirm this. Thus, it is of interest to include the 1B-Bangladesh SNP specific primers in future epidemiological studies, particularly in other (neighbouring) Asian countries such as India where genotype 1 is widely established (Monot et al., 2009).

In contrast to the general belief (Monot et al., 2009; Truman et al., 2011), we observed that subtype 1C does not form an independent subtype but actually belongs to subtype 1D. SNP61425 used to distinguish genotypes 1A-C is located at *esxA* encoding the virulence factor ESAT-6 (Monot et al., 2009). The Esx protein family also revealed high diversity in the more pathogenic mycobacterium, *M. tuberculosis* (Uplekar et al., 2011), and is involved in host-pathogen interaction. Of note is that ESAT-6 (ML0049) is a potent T-cell antigen (Geluk et al., 2002; Geluk et al., 2004), thus mutations in *esxA* gene might indicate drift due to immune pressure potentially explaining the occurrence of mutations at SNP61425 in different genotypes.

In a recent survey in 19 countries during 2009-2015 (Cambau et al., 2018), 8% of the cases presented mutations resulting in antimicrobial resistance and resistance to up to two different drugs was detected. In our study, which is the first investigating *M. leprae* drug resistance in Bangladesh, we detected no resistance by WGS, however, a partial missense mutation in the codon for Ser456 of the *rpoB* gene potentially leading to rifampicin resistance (n=1) was observed by Sanger sequencing. This could be the result of a mixed infection or an emerging mutation of the *M. leprae* strain occurring in the patient. Silent mutations in the *rpoB* gene were detected in several locations, which indicates that mutations do occur, and this may eventually lead to missense mutations conferring antimicrobial resistance. However, drug resistance is not only induced by genetic mutations in drug targets, efflux systems resulting in antimicrobial resistance have also been described for *M. leprae* (Machado et al., 2018). This mechanism of drug resistance is unnoticed in genomic tests and needs to be further investigated for leprosy especially in the light of the huge efforts recently initiated and WHO-endorsed for post-exposure prophylaxis (PEP) using antibiotic regimens (Barth-Jaeggi et al., 2016; Mieras et al., 2018; Richardus et al., 2019).

Despite our finding that NS samples were more frequently positive for *M. leprae* DNA, recovery of *M. leprae* whole genomes from SSS has proven to be more successful than from NS. This is due to the higher number of bacteria in SSS of patients. However, the importance of genotyping NS as well as skin biopsies or SSS to better understand transmission has been previously discussed (Fontes et al., 2017), as the nasal respiratory route remains one of the most plausible modes of infection (Bratschi et al., 2015; Araujo et al., 2016). In a recent study, skin biopsies and NS of patients were compared by VNTR typing and the authors found that out of 38 patients, differences between SSS and NS in seven loci were observed in 33 patients (Lima et al., 2016). Although the *M. leprae* genomes from SSS and NS analysed in our study were almost identical, we observed that genomes obtained from NS harboured more mutations, especially in previously reported (Benjak et al., 2018) hypermutated genes. This could be an indication of in-host evolution in the nasal mucosa, mixed infection or mixed colonization. Thus, it may imply that colonization occurred with two different strains causing a co-infection or that one is present, likely from a later colonization, but does not cause the disease.

The presence of mixed infections emphasises once more the importance of monitoring asymptomatic carriers, who may contribute to the spread of the pathogen. Therefore, providing PEP only to the (close) contacts of leprosy patients might not be sufficient to stop transmission. Instead, an approach including the entire community but targeting only individuals testing positive for *M. leprae* DNA or host immune markers associated to *M. leprae* infection, would represent a preferred strategy for PEP.

## Data Availability

Sequence data are available from the NCBI Sequence Read Archive (SRA) under the bioprojects PRJNA605605 and PRJNA592722, biosamples SAMN14072760-775 and SAMN13438761-771.

https://www.ncbi.nlm.nih.gov/bioproject/PRJNA605605

https://www.ncbi.nlm.nih.gov/bioproject/PRJNA592722

## Data availability

https://www.ncbi.nlm.nih.gov/bioproject/PRJNA605605

https://www.ncbi.nlm.nih.gov/bioproject/PRJNA592722

## Acknowledgements

The authors gratefully acknowledge all patients and control participants. LUMC. Erasmus MC and TLMI,B are part of the IDEAL (***I***nitiative for ***D***iagnostic and ***E***pidemiological ***A***ssays for ***L***eprosy) Consortium.

## Author contributions

Conceptualization: AG, MT, JHR

Data Curation: MT, JCR

Formal Analysis: MC, CA, AH, AB

Funding Acquisition: AG, JHR

Investigation: MT, CA, EV, LP, AH

Resources: MK, KA, PC, SC

Supervision: AG

Writing original draft: MT

Writing – Review & Editing: MT, AG

All authors reviewed, discussed, and agreed with manuscript.

## Supporting information captions

**Table S1. Cohort characterization**.

**Table S2. Primers and probes used in the study**.

**Table S3. *M. leprae* strain-specific SNPs**.

**Table S4. PCR primers for *M. leprae* strain-specific SNPs**.

**Table S5. WGS results**.

**Figure S1. Samples analysed by whole genome sequencing**.

**Supplementary Data 1. Overall subject and sample information, PCR, quantitative PCR (qPCR), genotyping and antimicrobial resistance results**.

**Supplementary Data 2. SNPs identified in 259 *M. leprae* genomes, including 27 genomes from this study**.

## References

(2017). “A guide for surveillance of antimicrobial resistance in leprosy”. (New Delhi: World Health Organization, Region Office for South-East Asia).

Araujo, S., Freitas, L.O., Goulart, L.R., and Goulart, I.M. (2016). Molecular evidence for the aerial route of infection of Mycobacterium leprae and the role of asymptomatic carriers in the persistence of leprosy. Clin Infect Dis 63(11), 1412–1420. doi: 10.1093/cid/ciw570.

Avanzi, C., Del-Pozo, J., Benjak, A., Stevenson, K., Simpson, V.R., Busso, P., et al. (2016). Red squirrels in the British Isles are infected with leprosy bacilli. Science 354(6313), 744–747. doi: 10.1126/science.aah3783.

Avanzi, C., Lecorché, E., Rakotomalala, F.A., Benjak, A., Rabenja, F.R., Ramarozatovo, L.S., et al. (In press). Population genomics of Mycobacterium leprae reveals a new genotype in Madagascar and Comoros. Front Microbiol.

Barbieri, R.R., Manta, F.S.N., Moreira, S.J.M., Sales, A.M., Nery, J.A.C., Nascimento, L.P.R., et al. (2019). Quantitative polymerase chain reaction in paucibacillary leprosy diagnosis: A follow-up study. PLoS Negl Trop Dis 13(3), e0007147. doi: 10.1371/journal.pntd.0007147.

Barth-Jaeggi, T., Steinmann, P., Mieras, L., van Brakel, W., Richardus, J.H., Tiwari, A., et al. (2016). Leprosy Post-Exposure Prophylaxis (LPEP) programme: study protocol for evaluating the feasibility and impact on case detection rates of contact tracing and single dose rifampicin. BMJ Open 6(11), e013633.

Benjak, A., Avanzi, C., Singh, P., Loiseau, C., Girma, S., Busso, P., et al. (2018). Phylogenomics and antimicrobial resistance of the leprosy bacillus Mycobacterium leprae. Nat Commun 9(1), 352. doi: 10.1038/s41467-017-02576-z.

Braet, S., Vandelannoote, K., Meehan, C.J., Brum Fontes, A.N., Hasker, E., Rosa, P.S., et al. (2018). The repetitive element RLEP is a highly specific target for detection of Mycobacterium leprae. J Clin Microbiol 56(3). doi: 10.1128/jcm.01924-17.

Bratschi, M.W., Steinmann, P., Wickenden, A., and Gillis, T.P. (2015). Current knowledge on Mycobacterium leprae transmission: a systematic literature review. Lepr Rev 86(2), 142–155.

Brito e Cabral, P., Junior, J.E., de Macedo, A.C., Alves, A.R., Goncalves, T.B., Brito e Cabral, T.C., et al. (2013). Anti-PGL1 salivary IgA/IgM, serum IgG/IgM, and nasal Mycobacterium leprae DNA in individuals with household contact with leprosy. Int J Infect Dis 17(11), e1005–1010. doi: 10.1016/j.ijid.2013.05.011.

Cambau, E., Saunderson, P., Matsuoka, M., Cole, S.T., Kai, M., Suffys, P., et al. (2018). Antimicrobial resistance in leprosy: results of the first prospective open survey conducted by a WHO surveillance network for the period 2009-15. Clin Microbiol Infect 24(12), 1305–1310. doi: 10.1016/j.cmi.2018.02.022.

Carvalho, R.S., Foschiani, I.M., Costa, M., Marta, S.N., and da Cunha Lopes Virmond, M. (2018). Early detection of M. leprae by qPCR in untreated patients and their contacts: results for nasal swab and palate mucosa scraping. Eur J Clin Microbiol Infect Dis 37(10), 1863–1867. doi: 10.1007/s10096-018-3320-9.

Cole, S.T., Eiglmeier, K., Parkhill, J., James, K.D., Thomson, N.R., Wheeler, P.R., et al. (2001). Massive gene decay in the leprosy bacillus. Nature 409(6823), 1007–1011.

Donoghue, H.D. (2019). Tuberculosis and leprosy associated with historical human population movements in Europe and beyond-an overview based on mycobacterial ancient DNA. Ann Hum Biol 46(2), 120–128. doi: 10.1080/03014460.2019.1624822.

Donoghue, H.D., Holton, J., and Spigelman, M. (2001). PCR primers that can detect low levels of Mycobacterium leprae DNA. J Med Microbiol 50(2), 177–182. doi: 10.1099/0022-1317-50-2-177.

Dwivedi, V.P., Banerjee, A., Das, I., Saha, A., Dutta, M., Bhardwaj, B., et al. (2019). Diet and nutrition: An important risk factor in leprosy. Microb Pathog 137, 103714. doi: 10.1016/j.micpath.2019.103714.

Fontes, A.N.B., Lima, L., Mota, R.M.S., Almeida, R.L.F., Pontes, M.A., Goncalves, H.S., et al. (2017). Genotyping of Mycobacterium leprae for better understanding of leprosy transmission in Fortaleza, Northeastern Brazil. PLoS Negl Trop Dis 11(12), e0006117. doi: 10.1371/journal.pntd.0006117.

Gama, R.S., Gomides, T.A.R., Gama, C.F.M., Moreira, S.J.M., de Neves Manta, F.S., de Oliveira, L.B.P., et al. (2018). High frequency of M. leprae DNA detection in asymptomatic household contacts. BMC Infect Dis 18(1), 153. doi: 10.1186/s12879-018-3056-2.

Gama, R.S., Souza, M.L.M., Sarno, E.N., Moraes, M.O., Goncalves, A., Stefani, M.M.A., et al. (2019). A novel integrated molecular and serological analysis method to predict new cases of leprosy amongst household contacts. PLoS Negl Trop Dis 13(6), e0007400. doi: 10.1371/journal.pntd.0007400.

Geluk, A., Duthie, M.S., and Spencer, J.S. (2011). Postgenomic Mycobacterium leprae antigens for cellular and serological diagnosis of M. leprae exposure, infection and leprosy disease. Leprosy review 82(4), 402–421.

Geluk, A., van Meijgaarden, K.E., Franken, K.L.M.C., Subronto, Y.W., Wieles, B., Arend, S.M., et al. (2002). Identification and characterization of the ESAT-6 homologue of Mycobacterium leprae and T-cell cross-reactivity with Mycobacterium tuberculosis. Infection and immunity 70(5), 2544–2548. doi: 10.1128/iai.70.5.2544-2548.2002.

Geluk, A., van Meijgaarden, K.E., Franken, K.L.M.C., Wieles, B., Arend, S.M., Faber, W.R., et al. (2004). Immunological crossreactivity of the Mycobacterium leprae CFP-10 with its homologue in Mycobacterium tuberculosis. Scandinavian journal of immunology 59(1), 66–70. doi: 10.1111/j.0300-9475.2004.01358.x.

Han, X.Y., Seo, Y.H., Sizer, K.C., Schoberle, T., May, G.S., Spencer, J.S., et al. (2008). A new Mycobacterium species causing diffuse lepromatous leprosy. Am J Clin Pathol 130(6), 856–864. doi: 10.1309/ajcpp72fjzzrrvmm.

Han, X.Y., and Silva, F.J. (2014). On the age of leprosy. PLoS neglected tropical diseases 8(2), e2544–e2544. doi: 10.1371/journal.pntd.0002544.

Honap, T.P., Pfister, L.A., Housman, G., Mills, S., Tarara, R.P., Suzuki, K., et al. (2018). Mycobacterium leprae genomes from naturally infected nonhuman primates. PLOS Neglected Tropical Diseases 12(1), e0006190. doi: 10.1371/journal.pntd.0006190.

Koboldt, D.C., Zhang, Q., Larson, D.E., Shen, D., McLellan, M.D., Lin, L., et al. (2012). VarScan 2: somatic mutation and copy number alteration discovery in cancer by exome sequencing. Genome Res 22(3), 568–576. doi: 10.1101/gr.129684.111.

Krause-Kyora, B., Nutsua, M., Boehme, L., Pierini, F., Pedersen, D.D., Kornell, S.-C., et al. (2018). Ancient DNA study reveals HLA susceptibility locus for leprosy in medieval Europeans. Nature communications 9(1), 1569–1569. doi: 10.1038/s41467-018-03857-x.

Kumar, B., Uprety, S., and Dogra, S. (2017). “Clinical diagnosis of leprosy,” in International textbook of leprosy, eds. D.M. Scollard & T.P. Gills. (www.internationaltextbookofleprosy.org.).

Lavania, M., Katoch, K., Katoch, V.M., Gupta, A.K., Chauhan, D.S., Sharma, R., et al. (2008). Detection of viable Mycobacterium leprae in soil samples: insights into possible sources of transmission of leprosy. Infect Genet Evol 8(5), 627–631. doi: 10.1016/j.meegid.2008.05.007.

Lavania, M., Katoch, K., Sachan, P., Dubey, A., Kapoor, S., Kashyap, M., et al. (2006). Detection of Mycobacterium leprae DNA from soil samples by PCR targeting RLEP sequences. J Commun Dis 38(3), 269–273.

Lima, L., Fontes, A.N.B., Li, W., Suffys, P.N., Vissa, V.D., Mota, R.M.S., et al. (2016). Intrapatient comparison of Mycobacterium leprae by VNTR analysis in nasal secretions and skin biopsy in a Brazilian leprosy endemic region. Lepr Rev 87(4), 486–500.

Machado, D., Lecorche, E., Mougari, F., Cambau, E., and Viveiros, M. (2018). Insights on Mycobacterium leprae Efflux Pumps and Their Implications in Drug Resistance and Virulence. Front Microbiol 9, 3072. doi: 10.3389/fmicb.2018.03072.

Manta, F.S.N., Barbieri, R.R., Moreira, S.J.M., Santos, P.T.S., Nery, J.A.C., Duppre, N.C., et al. (2019). Quantitative PCR for leprosy diagnosis and monitoring in household contacts: A follow-up study, 2011-2018. Sci Rep 9(1), 16675. doi: 10.1038/s41598-019-52640-5.

Martinez, A.N., Lahiri, R., Pittman, T.L., Scollard, D., Truman, R., Moraes, M.O., et al. (2009). Molecular determination of Mycobacterium leprae viability by use of real-time PCR. J Clin Microbiol 47(7), 2124–2130. doi: 10.1128/jcm.00512-09.

Martinez, A.N., Ribeiro-Alves, M., Sarno, E.N., and Moraes, M.O. (2011). Evaluation of qPCR-based assays for leprosy diagnosis directly in clinical specimens. PLoS Negl Trop Dis 5(10), e1354. doi: 10.1371/journal.pntd.0001354.

Mendum, T.A., Schuenemann, V.J., Roffey, S., Taylor, G.M., Wu, H., Singh, P., et al. (2014). Mycobacterium leprae genomes from a British medieval leprosy hospital: towards understanding an ancient epidemic. BMC Genomics 15, 270. doi: 10.1186/1471-2164-15-270.

Mieras, L.F., Taal, A.T., van Brakel, W.H., Cambau, E., Saunderson, P.R., Smith, W.C.S., et al. (2018). An enhanced regimen as post-exposure chemoprophylaxis for leprosy: PEP+. BMC Infect Dis 18(1), 506. doi: 10.1186/s12879-018-3402-4.

Mira, M.T., Alcais, A., Nguyen, V.T., Moraes, M.O., Di Flumeri, C., Vu, H.T., et al. (2004). Susceptibility to leprosy is associated with PARK2 and PACRG. Nature 427(6975), 636–640. doi: 10.1038/nature02326.

Moet, F.J., Meima, A., Oskam, L., and Richardus, J.H. (2004). Risk factors for the development of clinical leprosy among contacts, and their relevance for targeted interventions. Lepr Rev 75(4), 310–326.

Monot, M., Honore, N., Garnier, T., Araoz, R., Coppee, J.Y., Lacroix, C., et al. (2005). On the origin of leprosy. Science 308(5724), 1040–1042. doi: 10.1126/science/1109759.

Monot, M., Honore, N., Garnier, T., Zidane, N., Sherafi, D., Paniz-Mondolfi, A., et al. (2009). Comparative genomic and phylogeographic analysis of Mycobacterium leprae. Nat Genet 41(12), 1282–1289. doi: 10.1038/ng.477.

Penna, M.L., Penna, G.O., Iglesias, P.C., Natal, S., and Rodrigues, L.C. (2016). Anti-PGL-1 Positivity as a Risk Marker for the Development of Leprosy among Contacts of Leprosy Cases: Systematic Review and Meta-analysis. PLoS Negl Trop Dis 10(5), e0004703. doi: 10.1371/journal.pntd.0004703.

Richardus, R., Alam, K., Kundu, K., Chandra Roy, J., Zafar, T., Chowdhury, A.S., et al. (2019). Effectiveness of single-dose rifampicin after BCG vaccination to prevent leprosy in close contacts of patients with newly diagnosed leprosy: A cluster randomized controlled trial. Int J Infect Dis 88, 65–72. doi: 10.1016/j.ijid.2019.08.035.

Richardus, R.A., van der Zwet, K., van Hooij, A., Wilson, L., Oskam, L., Faber, R., et al. (2017). Longitudinal assessment of anti-PGL-I serology in contacts of leprosy patients in Bangladesh. PLOS Neglected Tropical Diseases 11(12), e0006083. doi: 10.1371/journal.pntd.0006083.

Ridley, D.S., and Jopling, W.H. (1966). Classification of leprosy according to immunity. A five-group system. Int. J. Lepr. Other Mycobact. Dis 34(3), 255–273.

Rimmer, A., Phan, H., Mathieson, I., Iqbal, Z., Twigg, S.R.F., Wilkie, A.O.M., et al. (2014). Integrating mapping-, assembly-and haplotype-based approaches for calling variants in clinical sequencing applications. Nat Genet 46(8), 912–918. doi: 10.1038/ng.3036.

Robinson, J.T., Thorvaldsdóttir, H., Winckler, W., Guttman, M., Lander, E.S., Getz, G., et al. (2011). Integrative genomics viewer. Nature biotechnology 29(1), 24–26. doi: 10.1038/nbt.1754.

Romero-Montoya, M., Beltran-Alzate, J.C., and Cardona-Castro, N. (2017). Evaluation and Monitoring of Mycobacterium leprae Transmission in Household Contacts of Patients with Hansen’s Disease in Colombia. PLoS Negl Trop Dis 11(1), e0005325. doi: 10.1371/journal.pntd.0005325.

Sales-Marques, C., Cardoso, C.C., Alvarado-Arnez, L.E., Illaramendi, X., Sales, A.M., Hacker, M.A., et al. (2017). Genetic polymorphisms of the IL6 and NOD2 genes are risk factors for inflammatory reactions in leprosy. PLoS Negl Trop Dis 11(7), e0005754. doi: 10.1371/journal.pntd.0005754.

Schilling, A.-K., Avanzi, C., Ulrich, R.G., Busso, P., Pisanu, B., Ferrari, N., et al. (2019a). British Red Squirrels Remain the Only Known Wild Rodent Host for Leprosy Bacilli. Frontiers in Veterinary Science 6(8). doi: 10.3389/fvets.2019.00008.

Schilling, A.-K., van Hooij, A., Corstjens, P., Lurz, P., DelPozo, J., Stevenson, K., et al. (2019b). Detection of humoral immunity to mycobacteria causing leprosy in Eurasian red squirrels (Sciurus vulgaris) using a quantitative rapid test.

Schuenemann, V.J., Avanzi, C., Krause-Kyora, B., Seitz, A., Herbig, A., Inskip, S., et al. (2018). Ancient genomes reveal a high diversity of Mycobacterium leprae in medieval Europe. PLoS Pathog 14(5), e1006997. doi: 10.1371/journal.ppat.1006997.

Schuenemann, V.J., Singh, P., Mendum, T.A., Krause-Kyora, B., Jager, G., Bos, K.I., et al. (2013). Genome-wide comparison of medieval and modern Mycobacterium leprae. Science 341(6142), 179–183. doi: 10.1126/science.1238286.

Sharma, R., Singh, P., Loughry, W.J., Lockhart, J.M., Inman, W.B., Duthie, M.S., et al. (2015). Zoonotic leprosy in the Southeastern United States. Emerg. Infect. Dis 21(12), 2127–2134. doi: 10.3201/eid2112.150501 [doi].

Singh, P., and Cole, S.T. (2011). Mycobacterium leprae: genes, pseudogenes and genetic diversity. Future Microbiol 6(1), 57–71. doi: 10.2217/fmb.10.153.

Spencer, J.S., and Brennan, P.J. (2011). The role of Mycobacterium leprae phenolic glycolipid I (PGL-I) in serodiagnosis and in the pathogenesis of leprosy. Lepr Rev 82(4), 344–357.

Suzuki, K., Takigawa, W., Tanigawa, K., Nakamura, K., Ishido, Y., Kawashima, A., et al. (2010). Detection of Mycobacterium leprae DNA from archaeological skeletal remains in Japan using whole genome amplification and polymerase chain reaction. PLoS One 5(8), e12422. doi: 10.1371/journal.pone.0012422.

Tio-Coma, M., Sprong, H., Kik, M., van Dissel, J.T., Han, X.Y., Pieters, T., et al. (2019). Lack of evidence for the presence of leprosy bacilli in red squirrels from North-West Europe. Transbound Emerg Dis. doi: 10.1111/tbed.13423.

Tió-Coma, M., Wijnands, T., Pierneef, L., Schilling, A.K., Alam, K., Roy, J.C., et al. (2019). Detection of Mycobacterium leprae DNA in soil: multiple needles in the haystack. Scientific Reports 9(1), 3165. doi: 10.1038/s41598-019-39746-6.

Truman, R.W., Andrews, P.K., Robbins, N.Y., Adams, L.B., Krahenbuhl, J.L., and Gillis, T.P. (2008). Enumeration of Mycobacterium leprae Using Real-Time PCR. PLOS Neglected Tropical Diseases 2(11), e328. doi: 10.1371/journal.pntd.0000328.

Truman, R.W., Singh, P., Sharma, R., Busso, P., Rougemont, J., Paniz-Mondolfi, A., et al. (2011). Probable zoonotic leprosy in the Southern United States. The New England journal of medicine 364(17), 1626–1633. doi: 10.1056/NEJMoa1010536.

Turankar, R.P., Lavania, M., Chaitanya, V.S., Sengupta, U., Darlong, J., Darlong, F., et al. (2014). Single nucleotide polymorphism-based molecular typing of M. leprae from multicase families of leprosy patients and their surroundings to understand the transmission of leprosy. Clin Microbiol Infect 20(3), O142–149. doi: 10.1111/1469-0691.12365.

Turankar, R.P., Lavania, M., Darlong, J., Siva Sai, K.S.R., Sengupta, U., and Jadhav, R.S. (2019). Survival of Mycobacterium leprae and association with Acanthamoeba from environmental samples in the inhabitant areas of active leprosy cases: A cross sectional study from endemic pockets of Purulia, West Bengal. Infect Genet Evol 72, 199–204. doi: 10.1016/j.meegid.2019.01.014.

Turankar, R.P., Lavania, M., Singh, M., Sengupta, U., Siva Sai, K., and Jadhav, R.S. (2016). Presence of viable Mycobacterium leprae in environmental specimens around houses of leprosy patients. Indian J Med Microbiol 34(3), 315–321. doi: 10.4103/0255-0857.188322.

Turankar, R.P., Lavania, M., Singh, M., Siva Sai, K.S., and Jadhav, R.S. (2012). Dynamics of Mycobacterium leprae transmission in environmental context: deciphering the role of environment as a potential reservoir. Infect Genet Evol 12(1), 121–126. doi: 10.1016/j.meegid.2011.10.023.

Uaska Sartori, P.V., Penna, G.O., Bührer-Sékula, S., Pontes, M.A.A., Gonçalves, H.S., Cruz, R., et al. (2020). Human Genetic Susceptibility of Leprosy Recurrence. Scientific reports 10(1), 1284–1284. doi: 10.1038/s41598-020-58079-3.

Uplekar, S., Heym, B., Friocourt, V., Rougemont, J., and Cole, S.T. (2011). Comparative genomics of Esx genes from clinical isolates of Mycobacterium tuberculosis provides evidence for gene conversion and epitope variation. Infection and immunity 79(10), 4042–4049. doi: 10.1128/IAI.05344-11.

van Beers, S.M., Izumi, S., Madjid, B., Maeda, Y., Day, R., and Klatser, P.R. (1994). An epidemiological study of leprosy infection by serology and polymerase chain reaction. Int J Lepr Other Mycobact Dis 62(1), 1–9.

Van Dissel, J.T., Pieters, T., Geluk, A., Maat, G., Menke, H.E., Tio-Coma, M., et al. (2019). Archival, paleopathological and aDNA-based techniques in leprosy research and the case of Father Petrus Donders at the Leprosarium ‘Batavia’, Suriname. Int J Paleopathol 27, 1–8. doi: 10.1016/j.ijpp.2019.08.001.

van Hooij, A., Tjon Kon Fat, E.M., Batista da Silva, M., Carvalho Bouth, R., Cunha Messias, A.C., Gobbo, A.R., et al. (2018). Evaluation of Immunodiagnostic Tests for Leprosy in Brazil, China and Ethiopia. Sci Rep 8(1), 17920. doi: 10.1038/s41598-018-36323-1.

van Hooij, A., Tjon Kon Fat, E.M., van den Eeden, S.J.F., Wilson, L., Batista da Silva, M., Salgado, C.G., et al. (2017). Field-friendly serological tests for determination of M. leprae-specific antibodies. Sci Rep 7(1), 8868. doi: 10.1038/s41598-017-07803-7.

van Hooij, A., van den Eeden, S., Richardus, R., Tjon Kon Fat, E., Wilson, L., Franken, K., et al. (2019). Application of new host biomarker profiles in quantitative point-of-care tests facilitates leprosy diagnosis in the field. EBioMedicine 47, 301–308. doi: 10.1016/j.ebiom.2019.08.009.

Wang, D., Xu, L., Lv, L., Su, L.Y., Fan, Y., Zhang, D.F., et al. (2015). Association of the LRRK2 genetic polymorphisms with leprosy in Han Chinese from Southwest China. Genes Immun 16(2), 112–119.

WHO (2019). Global leprosy update, 2018: moving towards a leprosy-free world. Weekly Epidemiological Record 94(35/36), 389–412.

Zhang, F.R., Huang, W., Chen, S.M., Sun, L.D., Liu, H., Li, Y., et al. (2009). Genomewide association study of leprosy. N Engl J Med 361(27), 2609–2618.

